# White matter integrity and its association with amyloid-PET and serum NfL in healthy *APOE4* homozygotes, heterozygotes and non-carries

**DOI:** 10.1101/2023.10.27.23297664

**Authors:** Claudia Tato-Fernández, Laura L. Ekblad, Elina Pietilä, Virva Saunavaara, Semi Helin, Riitta Parkkola, Henrik Zetterberg, Kaj Blennow, Juha O. Rinne, Anniina Snellman

**Affiliations:** Turku PET Centre, Turku University Hospital, Turku, Finland; Turku PET Centre, University of Turku, Turku, Finland; Department of Geriatric Medicine, Turku University Hospital, Turku, Finland; Department of Medical Physics, Division of Medical Imaging, Turku University Hospital, Finland; Department of Radiology, Turku University Hospital, Turku, Finland; Department of Radiology, University of Turku, Turku, Finland; Department of Psychiatry and Neurochemistry, Institute of Neuroscience and Physiology, the Sahlgrenska Academy at the University of Gothenburg, Mölndal, Sweden; Clinical Neurochemistry Laboratory, Sahlgrenska University Hospital, Mölndal, Sweden; Department of Neurodegenerative Disease, UCL Institute of Neurology, Queen Square, London, UK; UK Dementia Research Institute at UCL, London, UK; Hong Kong Center for Neurodegenerative Diseases, Clear Water Bay, Hong Kong, China; Wisconsin Alzheimer’s Disease Research Center, University of Wisconsin School of Medicine and Public Health, University of Wisconsin-Madison, Madison, WI, USA; Inst. of Neuroscience and Physiology, University of Gothenburg, Mölndal, Sweden; Clinical Neurochemistry Lab, Sahlgrenska University Hospital, Mölndal, Sweden; Paris Brain Institute, ICM, Pitié-Salpêtrière Hospital, Sorbonne University, Paris, France; Neurodegenerative Disorder Research Center, Division of Life Sciences and Medicine, and Department of Neurology, Institute on Aging and Brain Disorders, University of Science and Technology of China and First Affiliated Hospital of USTC, Hefei, P.R. China; InFLAMES Research Flagship, University of Turku, Turku, Finland

**Keywords:** Alzheimer’s disease, ApolipoproteinE, *APOE*, Magnetic resonance imaging, MRI, Diffusion tensor imaging, DTI, Neurofilament light chain, Positron emission tomography, [^11^C]PiB

## Abstract

Except for aging, *APOE ε4* allele (*APOE4*) is the most important risk factor for sporadic Alzheimer’s disease. *APOE4* carriers may have reduced capacity to recycle lipids, resulting in reduced white matter integrity. Here, we evaluated if white matter impairment measured by diffusion tensor imaging (DTI) differs between healthy individuals with different number of *APOE4* alleles and if white matter impairment associates with brain beta-amyloid (Aβ) load and serum levels of neurofilament light chain (NfL). We studied 96 participants (*APOE4/4*, *N* = 20; *APOE4/3*, *N* = 39; *APOE3/3*, *N* = 37; mean age 70.7 (SD 5.22) years, 63% females) with brain MRI including a DTI sequence (N = 96), Aβ-PET (N = 89) and a venous blood sample for serum NfL concentration measurement (N = 88). Fractional anisotropy (FA), mean diffusivity (MD), radial diffusivity (RD) and axial diffusivity (AxD) in six *a priori*-selected white matter regions-of-interest (ROIs) were compared between the groups using ANCOVA, with sex and age as covariates. A voxel-weighted average of FA, MD, RD and AxD was calculated for each subject, and correlations with Aβ-PET and NfL levels were evaluated. *APOE4/4* carriers exhibited higher MD and higher RD in the body of corpus callosum than *APOE4/3* (p = 0.0053 and p = 0.0049, respectively) and *APOE3/3* (p = 0.026 and p = 0.042). *APOE4/4* carriers had higher AxD than *APOE4/3* (p = 0.012) and *APOE3/3* (p = 0.040) in the right cingulum adjacent to cingulate cortex. In the total sample, composite MD, RD and AxD positively correlated with cortical Aβ load (r = 0.26 to 0.33, p < 0.013 for all) and with serum NfL concentrations (r = 0.31 to 0.36, p < 0.0028 for all). In conclusion, reduced white matter integrity was detected in cognitively unimpaired *APOE4/4* homozygotes compared to *APOE4/3* and *APOE3/3* carriers and reduced white matter integrity correlated with biomarkers of Alzheimer’s disease and neurodegeneration. White matter impairment seems to be an early phenomenon in the Alzheimer’s disease pathologic process in *APOE4/4* homozygotes.

**Highlights:**

- Cognitively normal elderly *APOE4* homozygotes showed impaired white matter integrity
- *APOE4* homozygotes showed higher diffusivity in corpus callosum and right cingulum
- MD, RD and AxD correlate with amyloid load assessed by PET in *APOE4* carriers
- MD, RD and AxD correlate with serum neurofilament light chain in *APOE4* carriers
- White matter impairment may be an early phenomenon in the AD pathologic process

## 1. Introduction

The ε4 allele for the *APOE* gene (*APOE4*) is the most important genetic risk factor for late-onset Alzheimer’s disease (AD) (Corder et al., 1993). The mechanism through which *APOE4* increases the risk for AD is not yet fully described, but the role of *APOE4* in lipid dysregulation has been recently highlighted (Blanchard et al., 2022). Apolipoprotein E (apoE) is involved in lipid transport in the brain (Espeseth et al., 2012), and lipids are essential components of the myelin sheath that encircles axons. *APOE4*-induced cholesterol dysregulations may decrease synaptogenesis and myelination (Bartzokis, 2011; Blanchard et al., 2022), which could contribute to the formation of toxic aggregates characteristic of AD, including beta-amyloid (Aβ) plaques (Lesser et al., 2011).

White matter impairment is part of the pathophysiological changes that an individual experience over the course of AD (Bronge et al., 2002; Ingelsson et al., 2004). Diffusion tensor imaging (DTI) is a well-established magnetic resonance imaging (MRI) technique to quantify the movement of water molecules in the brain (Basser et al., 1994), which can detect early neurodegeneration in patients of AD (Palesi et al., 2018). The tensor model characterizes diffusion with six parameters: three mutually orthogonal eigenvectors and their corresponding eigenvalues (O’Donnell & Westin, 2011). Fractional anisotropy (FA), a normalized variance of the eigenvalues (Basser & Pierpaoli, 1996; O’Donnell & Westin, 2011), describes the shape of the diffusion ellipsoid at every voxel. Mean diffusivity (MD) represents total diffusion in a voxel as an average of the eigenvalues (Basser & Pierpaoli, 1996). Measures of axial (AxD) and radial (RD) diffusivity describe diffusion along the axis of maximal apparent diffusion and its two orthogonal orientations in the perpendicular plane, respectively (O’Donnell & Westin, 2011). White matter pathology often causes anisotropy to decrease, which may be concomitant with subtle changes in one, or more, of the diffusion directions (Alexander et al., 2007).

Previous DTI studies have consistently reported damage to white matter tracts in patients with AD (Bachman et al., 2014; Esrael et al., 2021; Gallagher et al., 2023; Lim et al., 2012; Palesi et al., 2018; Racine et al., 2014). These findings have been linked to other risk factors for AD, including the presence of at least one copy of *APOE4* (Bagepally et al., 2012; Lee et al., 2016). However, studies with healthy *APOE4* carriers showed mixed results. Several DTI studies report degeneration of white matter tracts in healthy *APOE4* carriers (Bagepally et al., 2012; Cai et al., 2017; Cavedo et al., 2017; Douaud et al., 2011; Dowell et al., 2013; Gold et al., 2010; Heise et al., 2014; Nierenberg et al., 2005; Persson et al., 2006), but other investigations have been unable to replicate these findings (Adluru et al., 2014; Honea et al., 2009; Lyall et al., 2020; Nyberg & Salami, 2014; O’Dwyer et al., 2012; Westlye et al., 2012). Notably, these changes appear already in preclinical stages of AD (Gallagher et al., 2023).

Even though many studies have investigated the impact of *APOE* genotype on DTI metrics, the specific effects of *APOE4* homozygosity in healthy subjects are rarely analyzed. As only 25% of the Caucasian population carries the *APOE4* allele and prevalence of AD is elevated in this group (Gharbi-Meliani et al., 2021), it can be challenging to collect data from healthy elderly subjects who carry two copies of this allele. Most reports merge *APOE4/3* and *APOE4/4* when investigating the effects of *APOE4* in the brain. However, since *APOE4/4* carriers also have higher risk for vascular factors that could affect white matter integrity, including increased LDL cholesterol levels (Lesser et al., 2011) and white matter hyperintensities (WMHs) (Lyall et al., 2020), the changes detected with DTI could be expected to be more severe.

During recent years, it has become possible to investigate neuronal degeneration also with fluid-based biomarkers; however, little is still known about the association between DTI metrics and blood biomarkers of axonal degeneration in at-risk populations. Neurofilament light chain (NfL) is a protein expressed in neurons, which associates with other proteins to form the cytoskeleton of axons. Neurofilaments can be released in large quantities after axonal injury or degeneration (Schultz et al., 2020) and are nowadays measurable also from easily obtained blood samples (Gaetani et al., 2019). Elevated levels of serum NfL correlate with decreased FA and increased MD, AxD and RD, in patients of multiple sclerosis (Saraste et al., 2021) and AD (Schultz et al., 2020). To our knowledge, no previous study has investigated the relationship between serum NfL levels and DTI metrics in healthy *APOE4* carriers.

In this study, we aimed to expand previous DTI results by i) comparing DTI metrics of healthy elderly subjects with one, two, or no copies of the *APOE4* allele and ii) exploring their association with brain Aβ load assessed by [^11^C]PiB positron emission tomography (PET) and with serum NfL concentrations in the whole cognitively unimpaired sample. We hypothesized that *APOE4* carriers (*APOE4/4* and *APOE4/3*) would present impaired white matter integrity in a gene-dose dependent way when compared to *APOE3/3* carriers. We expected that impaired white matter integrity would associate with higher levels of serum NfL and Aβ pathology measured by PET.

## 2. Methods

### 2.1. Participants

A total of one hundred and nine healthy elderly adults (mini mental-state examination [MMSE] score ≥ 25, CERAD total score > 62 (Chandler et al., 2005), between the ages of 65 and 85 years old) from ASIC-E4 (Snellman et al., 2022) and CIRI-5Y (Ekblad et al., 2018) studies were included in this investigation. Data from both cohorts were acquired at Turku PET Centre. Main exclusion criteria were cognitive decline, neurological or psychiatric diseases, or contraindications for MRI and PET imaging. Participants from ASIC-E4 were recruited and *APOE* genotyped in collaboration with Auria Biobank (Turku, Finland) following a protocol already published (Snellman et al., 2022). Participants from CIRI-5Y were recruited from the Health2000 study population and *APOE* genotyped using the MassARRAY System (Sequenom, San Diego, CA) (Ekblad et al., 2018) with an adapted previously published method (Jänis et al., 2004). Both studies were approved by the Ethical Committee of the Hospital District of Southwest Finland. All participants signed a written informed consent according to the Declaration of Helsinki.

Six subjects were excluded from the initial sample because they were missing DTI data with reversed phase-encoding polarities. After examining DTI images, two subjects (*APOE3/3*) were excluded from analysis due to enlarged CSF spaces or incomplete head coverage. There were no subjects with *APOE2/4* genotype, but five subjects had *APOE2/3* genotype. Since DTI metrics of *APOE2/3* carriers were seen to significantly differ from those of *APOE3/3*, the genotypes could not be pooled together as a non-carrier group. Furthermore, given the small sample size of *APOE2* carriers (n = 5), we lacked sufficient statistical power to reach generalizable conclusions about this group. Thus, the main analyses and results are reported with the remaining ninety-six participants (Group 1: *APOE4/4*, *N* = 20; Group 2: *APOE4/3*, *N* = 39; Group 3: *APOE3/3*, *N* = 37). Supplementary table 1 and Supplementary figure 2 respectively show demographic and regional DTI results including *APOE2/3* carriers.

**Figure 1.**
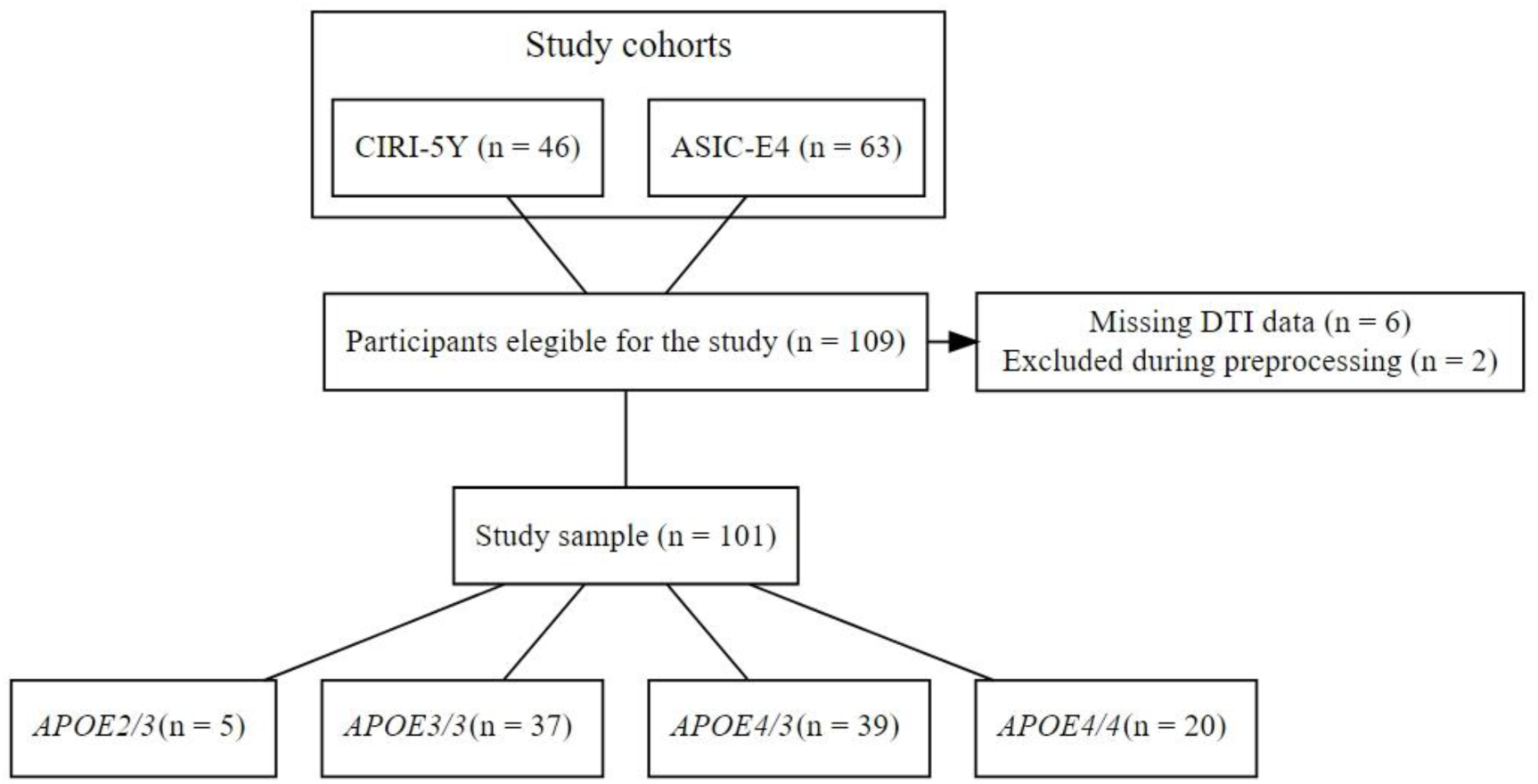
Study diagram. 109 participants were initially included from two cohorts, but eight subjects could not be included in the final study. *APOE2/3* carriers were not included in the main group analyses. DTI = diffusion tensor imaging

### 2.2. MRI Acquisition

Two scanners from the same manufacturer were used during data collection due to availability at the time of acquisition. 24 MRI scans were acquired with Philips Ingenia 3.0T systems with 20-channel dS head coil (Philips Healthcare, Amsterdam, the Netherlands) and the remaining 72 scans were acquired with Philips Ingenuity 3.0T TF PET-MR with 32-channel head coil (Philips Healthcare, Amsterdam, the Netherlands). In order to minimize possible confounding variation, the acquisition protocols were matched for both scanners. T1-weighted sequences, T2-weighted sequences, T2-weighted fluid-attenuated inversion recovery (FLAIR) sequences and DTI sequences were acquired for each participant. MRI images were reviewed by a neuroradiologist to exclude the presence of brain abnormalities.

The full imaging protocol has already been published (Snellman et al., 2022). DTI data were acquired using single-shot echo-planar imaging pulse sequences (TR = 6700 ms, TE = 120 ms, 2 x 2 x 2 mm voxels, 80 axial slices, slice thickness = 2 mm, no slice gap, field of view = 256 x 256 mm^2^, flip angle = 90°) with sensitivity encoding (SENSE) parallel imaging. One b=0 baseline volume was acquired with Philips Ingenia 3.0 T systems and four with Philips Ingenuity 3.0T PET-MR, along with 63 diffusion-weighted volumes with b-values of 1000 s/mm^2^. DTI sequences were followed by a single volume with b-value of 0, with the same acquisition parameters but opposing phase-encoding direction. A FLAIR image (TR = 8000 ms, TE = 337 ms, 1 x 1 x 1 mm voxels, field of view = 256 x 256 mm^2^, flip angle = 90°) was used to estimate computed Fazekas score using an automatic cNeuroimage analysis tool (Combinostics Oy, Tampere, Finland), with an adapted version of methods previously described (Koikkalainen et al., 2016).

### 2.3. PET Acquisition

Eighty-nine subjects additionally underwent PET imaging with the tracer [^11^C]PiB. PET images were acquired using an ECAT high-resolution research tomograph (HRRT, Siemens Medical Solutions, Knoxville, TN) with a spatial resolution of 2.5 mm. Data were acquired 40-90 minutes after injection of [^11^C]PiB (dose aimed at 500 MBq, minimum 250 MBq). PET scan duration was 50 minutes and it was followed by a 6 min transmission scan for attenuation correction, with a ^137^Cs point source. List-mode data was histogrammed into eight time frames (6 × 5 min; 2 × 10 min) and reconstructed with the 3D ordinary Poisson-ordered subset expectation maximization algorithm (OP-OSEM3D) with 16 subsets and 8 iterations and a voxel size of 1.22 × 1.22 × 1.22 mm.

### 2.4. Serum samples

Blood samples of eighty-eight participants were acquired in the morning after a 10-12-hour fasting period following in-house standard operating procedures (Snellman et al., 2022). Venous samples were analyzed at the Clinical Neurochemistry Laboratory of the University of Gothenburg (Mölndal, Sweden). The Single molecule array method and an HD-X analyzer (Quanterix) were used to measure serum NfL concentration (Simoa® NF-light™, #103186, Quanterix), following the instructions indicated by the manufacturer.

### 2.5. MRI processing

Image conversion from DICOM to NIFTI was done using dcm2niix v1.0.20190902 (Li et al., 2016). Slice-by-slice visual inspection was manually carried out for all volumes as quality control using FSLEyes’ Lightbox View. DTI volumes were corrected for subject motion, susceptibility- and eddy-current-induced distortions using the FMRIB Software Library v6.0.1 (Smith et al., 2004). Correction for susceptibility distortion was implemented using an adapted version of the “reverse gradient method” (Andersson et al., 2003). Outlier replacement and intra-volume movement correction were applied simultaneously with eddy current correction (Andersson et al., 2016; Andersson & Sotiropoulos, 2016). Non-brain tissue was removed using FSL’s brain extraction tool (Smith, 2002). Threshold for the brain mask was individually adjusted after visual inspection. The diffusion tensor model (Basser et al., 1994) was fit at each voxel to estimate parameter maps for FA, MD, AxD and RD.

The selection of ROIs was done separately from whole-brain analysis to mitigate potential bias. The following six regions were chosen based on previous literature researching the relationship between *APOE* gene and white matter integrity (Adluru et al., 2014; Cai et al., 2017; Cavedo et al., 2017; Gold et al., 2010; Racine et al., 2014): uncinate fasciculus (UF), genu, body and splenium of corpus callosum (G-CC, B-CC, S-CC), cingulum bundle projections to hippocampus (C-HC) and cingulum bundle running adjacent to the cingulate gyrus (C-CC) (Supplementary figure 1). UF, C-HC and C-CC were analyzed separately between hemispheres after testing that DTI metrics significantly differed contralaterally with paired-sample t-tests.

Participants’ FA, MD, AxD and RD maps were non-linearly transformed to MNI space using the FMRIB58_FA template as reference. FA maps were thresholded at 0.2 to correct for partial volume effects (PVEs). John Hopkins University (JHU) ICBM-DTI-81 atlas (Mori et al., 2008) was used to extract a brain mask of the selected ROIs. These masks were subsequently back-projected to each subject’s FA map in MNI152 space. Mean FA, MD, AxD and RD values were calculated by averaging the voxels within the boundaries of each ROI mask.

Individual FA maps were aligned to the template FMRIB58, in MNI152 space, using non-linear registration within the TBSS framework (Smith et al., 2006). A mean FA image was created and skeletonized. The tract skeleton was thresholded at a value of FA > 0.2. The non-linear warps obtained during FA image registration were subsequently applied to MD, AxD and RD maps.

### 2.6. PET processing

Analysis methods of Aβ PET data and *APOE4* gene dose-related differences in regional [^11^C]PiB binding have been previously described for a subset of participants (Snellman et al., 2023). Briefly, images were preprocessed using an automated pipeline (Karjalainen et al., 2020), which included co-registration to a T1-weighted MRI scan, ROI parcellation and PET kinetic modelling. Cerebellar grey matter was used as reference region. [^11^C]PiB binding was quantified as standardized uptake value ratios (SUVr) for the following ROIs: prefrontal cortex, parietal cortex, anterior cingulum, posterior cingulum, precuneus and lateral temporal cortex. These regions were averaged to obtain a volume-weighted composite (PiB-COMP) measure.

### 2.7. Statistical analyses

Main statistical analyses were performed using RStudio version 2022.12.0+353. For all numerical variables, normality was assessed with histograms, Shapiro-Wilk and Jarque-Bera tests. DTI metrics which did not fit the normal distribution were logarithmically transformed. Homoscedasticity was verified with Levene’s test.

Categorical demographic variables were compared between *APOE3/3, APOE4/3* and *APOE4/4* carriers with χ^2^ test. Numerical demographic variables were compared with ANOVA if normally distributed and Kruskal-Wallis test was used otherwise. Statistical significance was established at p < 0.05 (two-sided). If significant differences were found, Tukey’s honest significant difference (HSD) or Dunn’s test were used to determine which specific groups differed significantly from each other.

FA, MD, AxD and RD at each of the ROIs were compared between *APOE3/3, APOE4/3* and *APOE4/4* carriers with one-way ANCOVA, with age and sex as covariates of no interest. If a significant difference was found, Tukey’s HSD was used to determine the direction of differences and Cohen’s d was used to assess the effect size. To test that our results were independent of the scanner used at the time of acquisition and of WMHs, measured as computed Fazekas score, we included these two variables as covariates in a second model.

By conducting ROI analysis, we aimed to quantify white matter integrity in a controlled number of regions relevant to the development AD, based on predefined hypotheses. Moreover, the main purpose of ASIC-E4 and CIRI-5Y cohorts was not to assess differences in white matter integrity, which makes this study exploratory in nature. This means it may lack sufficient statistical power to detect subtle differences, thus providing false negatives. For these reasons, following the guidelines of statistical theory (Rothman, 1990) and current research (Lyall et al., 2020; Svärd et al., 2017), uncorrected alpha < 0.05 was considered nominally significant in our regional analyses. We additionally applied false discovery rate (FDR) correction to our findings (Benjamini & Hochberg, 1995), but results are reported without correction for multiple comparisons unless stated otherwise.

To evaluate the association of DTI measures with serum NfL concentration and global Aβ deposition estimated by PiB-COMP, mean scores of FA, MD, AxD and RD were calculated by computing the voxel-weighted average of our ROIs. Spearman’s rank-order test was used to assess the correlation coefficient in the whole sample and stratified by *APOE* status, which was considered significant at p < 0.05.

Voxel-wise differences were assessed with the GLM design using the Randomise tool in FSL (Winkler et al., 2014). DTI metrics were compared between *APOE4/4*, *APOE4/3* and *APOE3/3* carriers using one-way ANCOVA, including mean-centered age and sex as covariates of no interest. The number of permutations was set to 5000 based on the threshold-free cluster enhancement (TFCE) (Smith & Nichols, 2009). Results were considered significant at p < 0.05 with the family wise error (FWE) correction for multiple comparisons. The resultant clusters were labelled according to JHU ICBM-DTI-81 white matter labels atlas.

## 3. Results

### 3.1. Study population

Table 1 shows the distribution of demographic variables stratified by *APOE* genotype. Our sample included 96 participants (mean age = 70.7 SD = 5.22), of whom 63% were females. All groups were well-matched for age (p = 0.29), sex (p = 0.89), educational level (p = 0.68) and body-mass index (p = 0.16). There were significant differences in [^11^C]PiB SUVrs between *APOE4/4*, *APOE4/3* and *APOE3/3* carriers (p = 0.0010). *APOE3/3* showed significantly lower Aβ deposition than *APOE4/4* (p = 0.0013) and *APOE4/3* (p = 0.017).

**Table 1.**
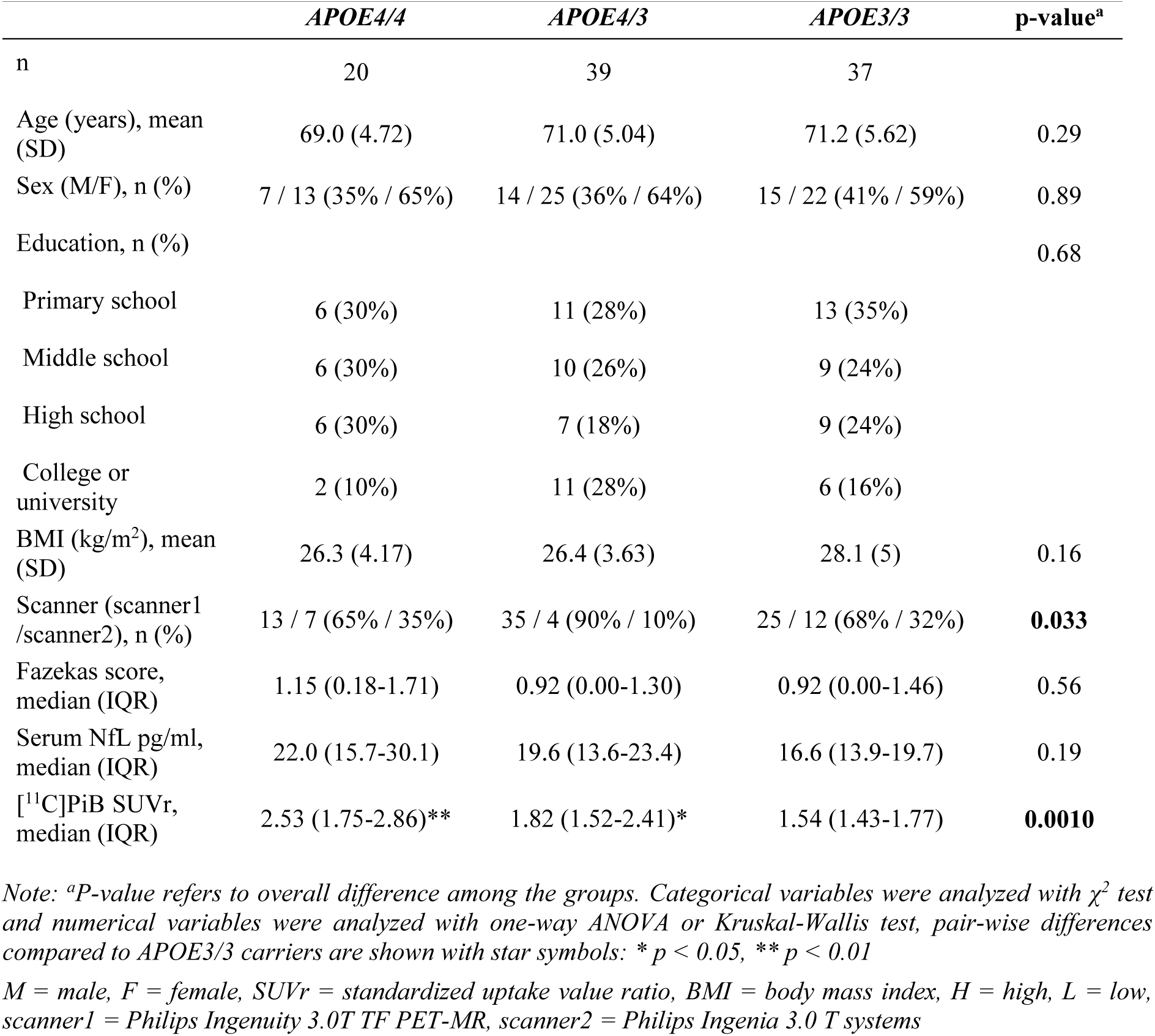
Demographic variables.

|  | *APOE4/4* | *APOE4/3* | *APOE3/3* | p-value<sup>a</sup> |
| --- | --- | --- | --- | --- |
| n | 20 | 39 | 37 |  |
| Age (years), mean (SD) | 69.0 (4.72) | 71.0 (5.04) | 71.2 (5.62) | 0.29 |
| Sex (M/F), n (%) | 7 / 13 (35% / 65%) | 14 / 25 (36% / 64%) | 15 / 22 (41% / 59%) | 0.89 |
| Education, n (%) |  |  |  | 0.68 |
| Primary school | 6 (30%) | 11 (28%) | 13 (35%) |  |
| Middle school | 6 (30%) | 10 (26%) | 9 (24%) |  |
| High school | 6 (30%) | 7 (18%) | 9 (24%) |  |
| College or university | 2 (10%) | 11 (28%) | 6 (16%) |  |
| BMI (kg/m<sup>2</sup>), mean (SD) | 26.3 (4.17) | 26.4 (3.63) | 28.1 (5) | 0.16 |
| Scanner (scanner1 / scanner2), n (%) | 13 / 7 (65% / 35%) | 35 / 4 (90% / 10%) | 25 / 12 (68% / 32%) | **0.033** |
| Fazekas score, median (IQR) | 1.15 (0.18-1.71) | 0.92 (0.00-1.30) | 0.92 (0.00-1.46) | 0.56 |
| Serum NfL pg/ml, median (IQR) | 22.0 (15.7-30.1) | 19.6 (13.6-23.4) | 16.6 (13.9-19.7) | 0.19 |
| [$^{11}\text{C}$]PiB SUVr, median (IQR) | 2.53 (1.75-2.86)** | 1.82 (1.52-2.41)* | 1.54 (1.43-1.77) | **0.0010** |

Note: <sup>a</sup>P-value refers to overall difference among the groups. Categorical variables were analyzed with $\chi^2$ test and numerical variables were analyzed with one-way ANOVA or Kruskal-Wallis test, pair-wise differences compared to *APOE3/3* carriers are shown with star symbols: \* $p < 0.05$ , \*\* $p < 0.01$
M = male, F = female, SUVr = standardized uptake value ratio, BMI = body mass index, H = high, L = low, scanner1 = Philips Ingenuity 3.0T TF PET-MR, scanner2 = Philips Ingenia 3.0 T systems

### 3.2. Regional analysis

DTI variables for the *APOE4/4, APOE4/3* and *APOE3/3* groups are presented in Figure 2A-D. First, there were no differences in FA between *APOE4/4, APOE4/3* and *APOE3/3* in any of the chosen ROIs (all p > 0.13, ANCOVA, Fig 2A), whereas *APOE4/4* showed increased MD in the B-CC (F = 3.37, p = 0.039, ANCOVA) when compared to *APOE4/3* (p = 0.0053, Cohen’s d = 0.68) and *APOE3/3* (p = 0.026, Cohen’s d = 0.45) (Fig 2B). In this region, *APOE4/4* also exhibited higher RD (F = 3.40, p = 0.038, ANCOVA) than *APOE4/3* (p = 0.0049, Cohen’s d = 0.69) and *APOE3/3* (p = 0.042, Cohen’s d = 0.39) (Fig 2C). A significant increase in AxD was also found in the RC-CG (F = 3.94, p = 0.023, ANCOVA) when comparing *APOE4/4* against *APOE4/3* (p = 0.012, Cohen’s d = 0.41) and *APOE3/3* (p = 0.040, Cohen’s d = 0.39) (Fig 2D). No significant differences in regional MD, AxD nor RD were found between *APOE4/3* and *APOE3/3*. All of the results remained significant after adjusting the analysis for the type of MRI scanner used during image acquisition (all p < 0.040) and number of WMHs (all p < 0.035). However, no significant differences remained after FDR correction (all fully adjusted p > 0.21).

**Figure 2.**
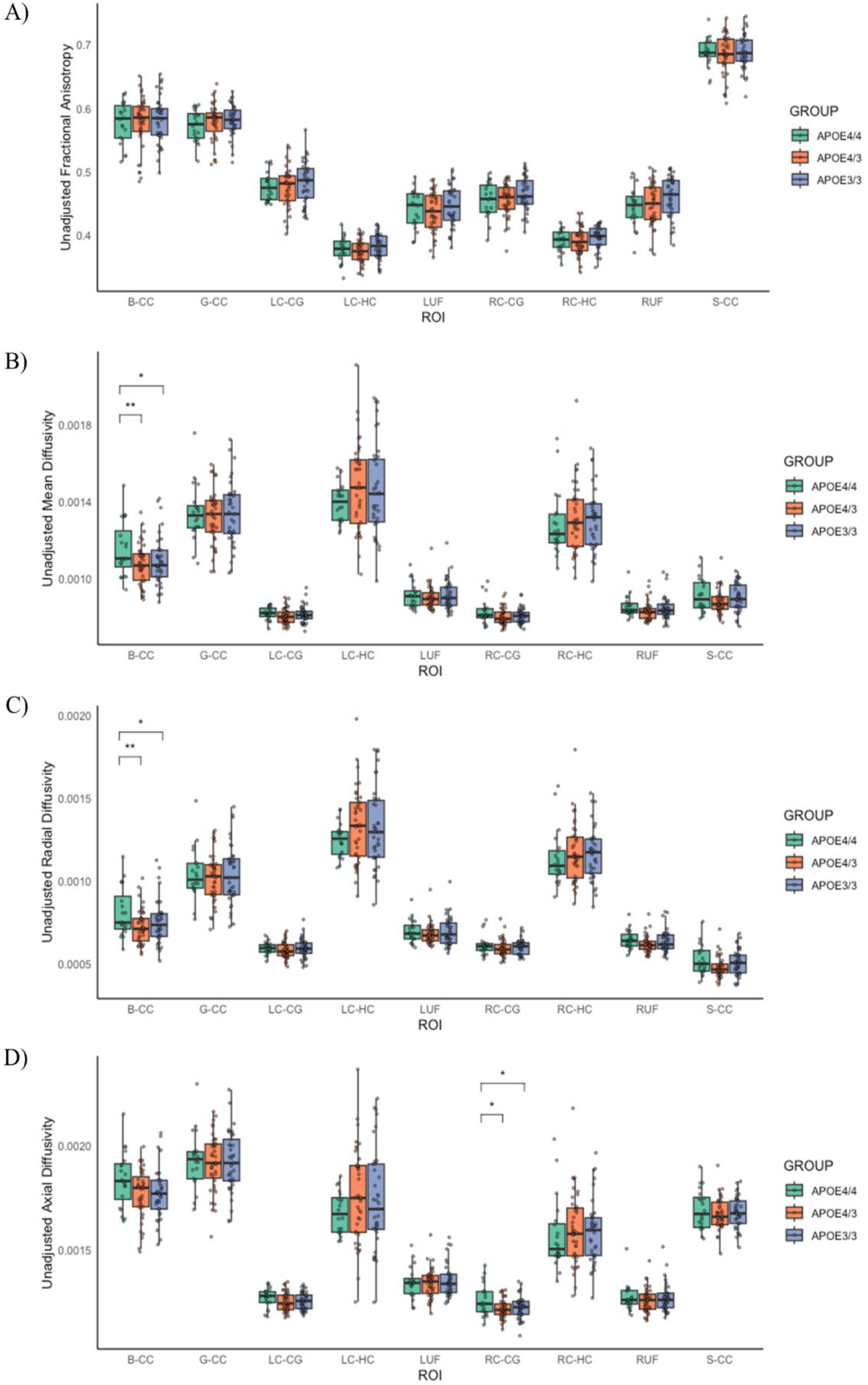
Regional differences in fractional anisotropy (2A), mean diffusivity (2B), radial diffusivity (2C) and axial diffusivity (2D) between *APOE4/4*, *APOE4/3* and *APOE3/3* estimated by ANCOVA, corrected for age and sex. Pair-wise differences in Tukey’s HSD test are shown with star symbols: *\* p < 0.05, ** p < 0.01* B-CC = body of corpus callosum, G-CC = genu of corpus callosum, LC-CG = left cingulum adjacent to cingulate gyrus, LC-HC = left cingulum adjacent to hippocampus, LUF = left uncinate fasciculus, RC-CG = right cingulum adjacent to cingulate gyrus, RC-HC = right cingulum adjacent to hippocampus, RUF = right uncinate fasciculus, S-CC = splenium of corpus callosum

Group comparisons including *APOE2/3* carriers (n = 5) are shown in Supplementary Figure 2. *APOE2/3* carriers showed increased MD and RD when compared to *APOE4/3*. They also exhibited higher AxD than *APOE4/3* and *APOE3/3* carriers.

### 3.3. Whole-brain analysis

Results obtained from the whole-brain analysis are presented in Figure 3. First, we did not find any significant differences between *APOE4/4, APOE4/3* and *APOE3/3* groups (all p > 0.09, FWE-corrected). However, an exploratory analysis with a more liberal threshold (uncorrected p < 0.001) revealed subtle differences in MD and AxD between *APOE4/4*, *APOE4/3* and *APOE3/3* (Fig. 3). The remaining contrasts showed that *APOE4/4* carriers exhibited higher MD than *APOE4/3* and *APOE3/3* carriers, in line with the ROI-level findings. In addition, *APOE4/3* carriers showed greater AxD than *APOE3/3*. The contrasts corresponding to individual t-tests between the three groups are displayed in Supplementary figure 2A-B.

**Figure 3.**
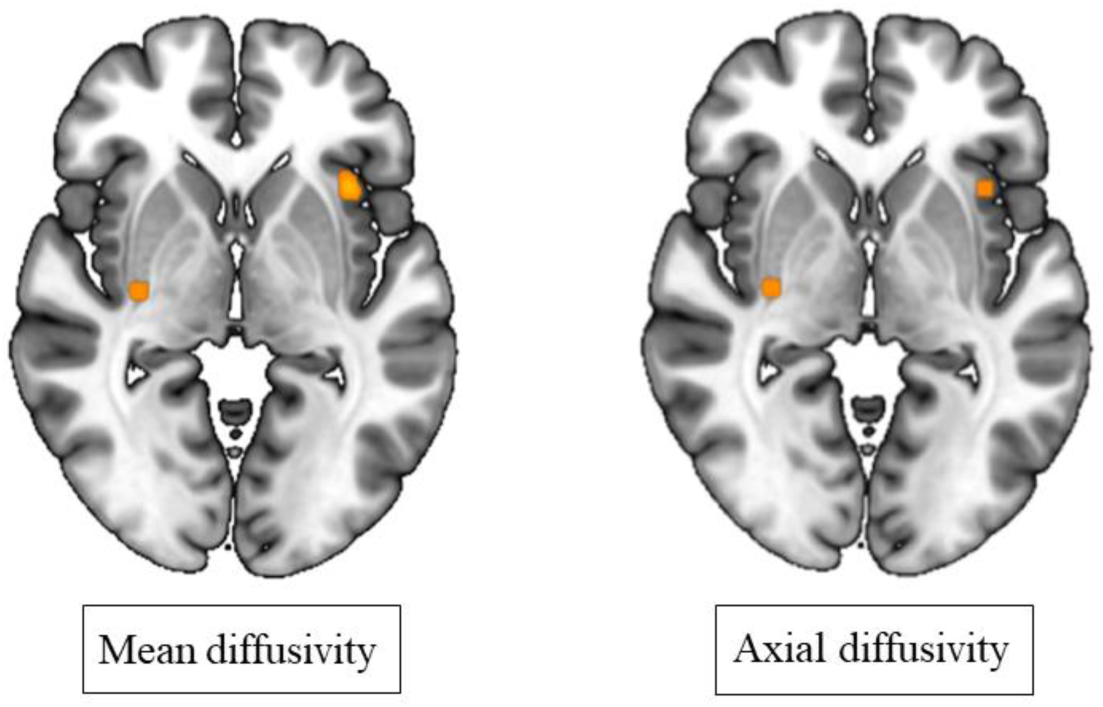
Comparing whole-brain white matter integrity between *APOE4/4*, *APOE4/3* and *APOE3/3* with TBSS. Subthreshold clusters (shown in orange) where the three groups differed in mean and axial diffusivity were found in exploratory analyses (p < 0.001, corrected for age and sex, uncorrected for multiple comparisons). Clusters have been filled with “tbss fill” in FSL, for visualization purposes.

### 3.4. Association between DTI parameters and serum NfL concentrations

We found that, in our cognitively unimpaired sample, higher serum NfL levels were associated with higher MD (r = 0.33 p = 0.0019), RD (r = 0.36, p = 0.00061) and AxD (r = 0.31, p = 0.0028) (Fig. 4). On the contrary, no significant association was detected with FA. When stratified by *APOE* group, we did not find any significant correlation between DTI scalars and serum NfL levels in *APOE4* non-carriers (all p > 0.25). The correlation between MD and serum NfL concentrations was mostly driven by *APOE4/3* carriers (r = 0.55, p = 0.00083) and, to a lesser extent, by *APOE4/4* carriers (r = 0.48, p = 0.034). RD similarly correlated with serum NfL in *APOE4/3* carriers (r = 0.55, p = 0.00085) and *APOE4/4* carriers (r = 0.5, p = 0.023). The correlation between AxD and serum NfL levels was solely driven by *APOE4/3* carriers (r = 0.5, p = 0.0026). *APOE4/4* carriers showed a trend towards a positive correlation which did not reach significance (r = 0.39, p = 0.089).

**Figure 4.**
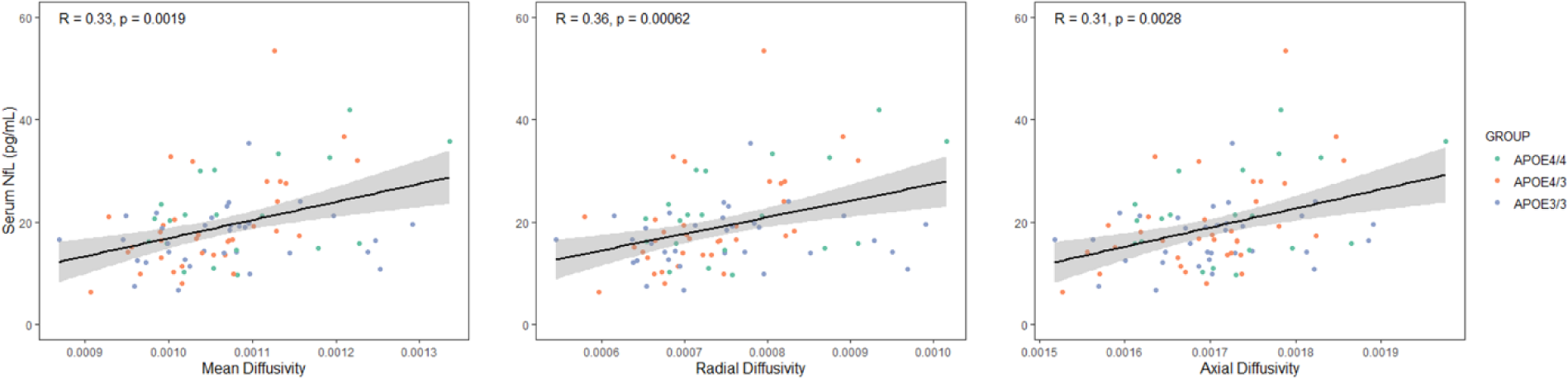
Spearman’s correlation coefficients between DTI metrics and serum NfL concentrations. High serum levels of NfL positively correlate with mean, radial and axial diffusivity in the whole sample, but not in subjects who do not carry any *APOE4* alleles NfL = neurofilament light chain

### 3.5. Association between DTI parameters and brain Aβ load

High brain Aβ load was associated with higher MD (r = 0.29, p = 0.0063), RD (r = 0.26, p = 0.013) and AxD (r = 0.33, p = 0.013) (Fig. 5). Again, no significant correlations were found for FA. Stratification by *APOE* genotype revealed no significant correlations between DTI scalars and brain Aβ load in non-carriers (all p > 0.18). In the *APOE4/4* group, only AxD significantly correlated with Aβ load (r = 0.53, p = 0.02), whereas the correlation with MD was borderline significant (r = 0.44, p = 0.058). The remaining correlations were driven by *APOE4/3* carriers (all p < 0.015).

**Figure 5.**
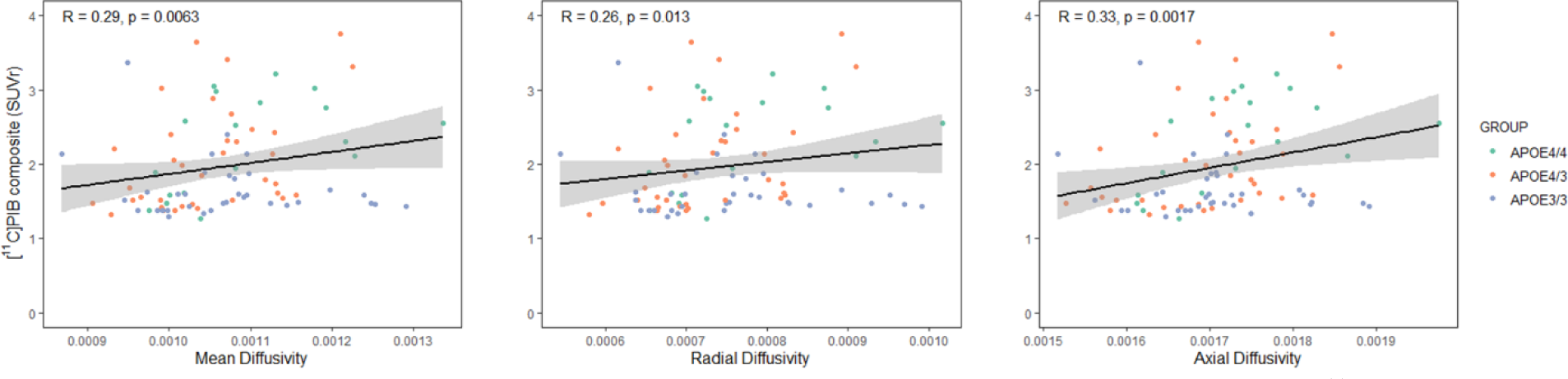
Spearman’s correlation coefficients between DTI metrics and Aβ deposition. High [^11^C]PiB uptake positively correlates with mean, radial and axial diffusivity in the whole sample, but not in subjects who do not carry any *APOE4* alleles

## 4. Discussion

We aimed to test the differential effects of *APOE4* hetero- and homozygosity on white matter integrity in elderly healthy individuals (n = 96). We demonstrate that white matter integrity was reduced in *APOE4/4* homozygotes when compared to *APOE4/3* and *APOE3/3* carries, and that indicators of white matter impairment correlated with biomarkers of AD and neurodegeneration in this cognitively well-preserved population. Recent research has resurfaced the hypothesis that *APOE4* allele impairs white matter integrity (Blanchard et al., 2022; Lee et al., 2022). Our results moderately supported our initial hypothesis, stating that *APOE4* allele would damage white matter integrity already in healthy elderly carriers, and particularly in *APOE4* homozygotes.

We found increased MD and RD in the body of corpus callosum in *APOE4/4* carriers, compared to *APOE4/3* and *APOE3/3* groups. *APOE4/4* also exhibited higher AxD than *APOE4/3* and *APOE3/3* carriers in the right cingulum adjacent to the cingulate gyrus. Although we found significant differences in MD, RD and AxD in certain white matter regions, we did not find any differences in FA between our three groups. These findings are in line with previous DTI studies where no associations between *APOE4* allele and FA have been found (Dowell et al., 2013; Honea et al., 2009; Nyberg & Salami, 2014; Rieckmann et al., 2016; Westlye et al., 2012). One of these studies also found increased AxD in *APOE4* carriers, proving that separate eigenvalue analysis offers specificity and additional perspectives (Dowell et al., 2013). MD is sensitive to tissue necrosis, as RD tends to be to myelin lesions, and AxD can measure axonal fiber coherence (Alexander et al., 2007). Our findings point that *APOE4/4* carriers may present moderate axonal damage that is unlikely to be due to age-related changes. A previous DTI study with few *APOE4/4* subjects (n = 10) did not report differences between *APOE4* homo- and heterozygotes (Persson et al., 2006), but we expected that subtle differences would become noticeable when sample size increases. The disparity of previous results may be due to heterogeneities in cohorts and regions analyzes. The first DTI studies which reported *APOE4-*related changes were conducted with different methodologies (Nierenberg et al., 2005; Persson et al., 2006). Atlas-based ROI analysis facilitates reproducibility and reduces researcher bias, compared to manual delimitation.

We did not find the gene-dose effects we expected, because *APOE4/3* and *APOE3/3* carriers did not significantly differ from each other at regional level, probably related to the limited numbers in each group. DTI metrics are not specific indicators of AD. Thus, there could also be other variables mediating these findings. Even though the novelty of our results is due to the recruitment of a healthy elderly *APOE4/4* group, there might be a bias in these participants. Since most *APOE4/4* homozygotes develop AD, it has been noted that these “healthy survivors” are exceptional and their white matter tracts are preserved in a way that might not represent their population (Thompson et al., 2011; Westlye et al., 2012). Similarly, *APOE4/3* carriers who are still cognitively healthy at approximately 70 years probably have only very subtle AD-related changes in their brain. This is a plausible explanation of why *APOE4/3* did not show significantly impaired white matter integrity against *APOE3/3* carriers in this relatively small study population.

Differences in DTI scalar measures were not significant at whole-brain level when we compared the three *APOE4* groups to each other (all FWE-corrected p > 0.09), however, few small significant clusters appeared when we used a more liberal threshold (uncorrected p < 0.001). We tested the direction of these differences and found that *APOE4/4* showed increased MD compared to *APOE4/3* and *APOE3/3*, and *APOE4/3* exhibited higher AxD than *APOE3/3* (FWE-corrected p < 0.05), thus aligning to our ROI analysis. TBSS was run after our primary analysis to confirm our findings. It is possible that whole-brain ANCOVA did not reach significant threshold because the ROI approach focused only on regions vulnerable to AD pathology, whereas analysis of large brain maps included a whole white matter skeleton.

We chose a set of ROIs (B-CC, G-CC, S-CC, LC-CG, RC-CG, LC-CH, RC-CH, RUF, LUF) for regional analysis aimed to specifically test the effects of early AD, since *APOE4* carriers in a subset of our cohort already exhibited high Aβ loads (Snellman et al., 2023). We found significant white matter impairment in the cingulum and corpus callosum of *APOE4* homozygotes. Additional studies showed that these regions are damaged in patients of AD (Esrael et al., 2021; Gallagher et al., 2023; Lim et al., 2012; Palesi et al., 2018), and already in healthy *APOE4* heterozygotes (Adluru et al., 2014; Cai et al., 2017). AD affects the entorhinal cortex and the hippocampus at early stages (Braak & Braak, 1991). The cingulum connects components of the limbic system, including the entorhinal cortex, and the corpus callosum interconnects the cerebral hemispheres. These are networks involved in higher cognitive functions. Even though our participants did not exhibit cognitive decline, microstructural axonal damage might precede the onset of symptoms, and thus becomes an important feature in early detection. In addition, the fornix is a region of particular interest in the study of AD. It is a major output tract from the hippocampus that is thought to be involved in episodic memory. Damage to the fornix has been consistently reported in studies of patients with dementia (Aggleton et al., 2016; Oishi & Lyketsos, 2014), so we initially selected this region for our study. Despite its relevance from the biological perspective, the fornix is susceptible to PVEs, given its proximity to cerebrospinal fluid (Oishi & Lyketsos, 2014; Rieckmann et al., 2016). Although FA is quantified between 0 and 1, FA values below 0.20 are unlikely to represent white matter tracts (Rieckmann et al., 2016; Smith et al., 2006). 30.7% of our subjects exhibited values of FA < 0.2 in the fornix before thresholding, while FA was consistently higher in the remaining regions. Thus, the fornix was not included as ROI in our final analyses.

In our statistical analyses, we controlled for possible nuisance covariates – including age, sex, type of scanner used during acquisition and Fazekas scores. WMHs are regions of increased signal intensity on T2-weighted MRI, which might be due to damage to blood vessels and inflammation. They are commonly found in elderly subjects, and they are related to cognitive decline and an increased risk for dementia (De Groot et al., 2001; Tubi et al., 2020). AD and *APOE4* allele have been previously associated with WMHs (Lyall et al., 2020; Palesi et al., 2018), although Fazekas score did not significantly vary within our cohort. Our results remained significant when we adjusted our analyses for WMHs. Including WMHs and type of scanner as covariates did not improve the explanatory value of the models (adjusted R^2^ increased on average 0.03 after including these two variables), hence we decided to only include sex and age as covariates in our main results, to avoid overadjustment.

The differences in MD, RD and AxD which we found at regional level were attenuated when we implemented FDR correction. The effect sizes of our results were mostly small, but we found medium effects of *APOE4* homozygosity in the body of corpus callosum (Cohen’s d > 0.50). As it has been previously noted (Cox et al., 2016; Lyall et al., 2020; Rothman, 1990), adjusting for type-1 error when analyzing brain MRI phenotypes might be overly cautious, and it is more reliable to test the replicability of findings with different cohorts.

### 4.1. *APOE2* allele and white matter integrity

The *APOE2* allele is generally considered to be neuroprotective (Nagy et al., 1995), but its effects on white matter integrity have rarely been investigated, and with inconsistent results (Chiang et al., 2012; Lyall et al., 2014). Despite this mixed evidence, we expected the *APOE2* allele to protect white matter tracts, but our findings indicated the opposite. Our sample of *APOE2/3* carriers was advanced in age, which means they may have developed non-AD related pathologies that could affect DTI measures. However, since our *APOE2* carrier group only had five subjects, we cannot provide conclusive insights on this topic.

### 4.2. DTI and serum NfL

NfL is a measure of the intensity of axonal degeneration, measurable in cerebrospinal fluid, and nowadays also in blood samples (Gaetani et al., 2019). As our secondary objective, we aimed to expand current literature by relating our findings in DTI with serum NfL levels in cognitively unimpaired at-risk individuals. We hypothesized that serum NfL levels in *APOE4* carriers would be directly related to MD, RD, AxD and inversely related to FA, as it has been found in patients of multiple sclerosis (Saraste et al., 2021) and AD (Schultz et al., 2020). We found no correlations between FA and serum NfL levels, but diffusivity scores (MD, RD and AxD) in regions vulnerable to AD pathology correlated with serum NfL in the expected direction, where increased NfL levels were associated with increased diffusivity scores in the whole sample, but not in *APOE3/3* carriers, although these correlations were more prominent in subjects carrying only one *APOE4* allele. The associations found in the present study highlight the value of serum NfL as a biomarker for axonal degeneration neurological diseases. In our sample, serum NfL levels and DTI metrics support each other, therefore confirming their potential to detect white matter impairment in subjects at genetic risk for AD.

### 4.3. DTI and Aβ load

Plaques of Aβ are amongst the hallmark pathological findings of AD. A reduced capacity to clear Aβ plaques in the extracellular space is one of the negative effects that *APOE4* allele is suggested to induce (Husain et al., 2021). Aβ accelerates the rate at which FA declines in longitudinal studies (Rieckmann et al., 2016). Elderly cognitively normal *APOE4* carriers present increased Aβ load, in a gene-dose dependent way (Snellman et al., 2023). The relationship between Aβ and DTI differs across cohorts and regions (Racine et al., 2014; Wang et al., 2020), presumably due to model constraints, since AD affects regions with heterogeneous fiber orientations (Douaud et al., 2011).

The finding that MD, RD and AxD in white matter bundles relevant for AD are correlated with regional [^11^C]PiB binding could sign that white matter damage in subjects at genetic risk for AD is a byproduct of large concentrations of Aβ in the brain. *APOE3/3* and *APOE4/3* carriers in our cohort had lower Aβ load than *APOE4/4* and their white matter tracts were less damaged. Numerous studies have failed to link *APOE* gene and early white matter impairment, thus supporting the idea that white matter abnormalities are more related to AD pathology than *APOE* genotype. However, it is not possible to deduce from these findings whether Aβ harms white matter integrity, or both increased diffusivity and Aβ load are a consequence of the pathology the brain experiences in AD.

### 4.4. Strengths and limitations

A major strength of this project is the recruitment of 20 healthy *APOE4/4* carriers who underwent a protocol including MRI, PET and serum samples. Thanks to the collaboration with Auria biobank during recruitment, it was possible to include a sample of *APOE4/4* carriers large enough to conduct separate group analysis, therefore improving statistical power.

This study also has several limitations. DTI is an indirect measure, unable to accurately measure crossing fibers, and its outputs cannot be considered specific markers of AD. Nonetheless, the DTI model was well suited to our single-shell diffusion data in order to conduct group comparisons. We used two scanners during image acquisition, with slightly different protocols, and uneven distributions among groups. This introduced an additional confound to our analyses, although it did not influence our results.

Other limitations are related to our sample. Firstly, our study cohort did not include a large enough proportion of *APOE2/3* carriers to analyze these participants as a separate group. Secondly, the age range of our healthy *APOE4/4* carriers was slightly younger when compared to the other groups, although they did not significantly differ in statistical analyses (p = 0.29, ANOVA). The eldest *APOE4/4* carrier in our cohort was 76 years old, while our non-carrier group included participants up to 86 years old, who might have developed other pathologies such as vascular diseases. DTI findings did not correlate with AD biomarkers in this *APOE* non-carrier group, which supports the theory that white matter degeneration is a sign of early AD in subjects at genetic risk for the disease, but not in non-*APOE4* carriers.

## 5. Conclusions

Our main finding was that healthy elderly *APOE4/4* carriers showed significantly increased MD, RD and AxD compared to *APOE4/3* and *APOE3/3* carriers in regions vulnerable to AD. These indicators positively correlated with Aβ deposition and serum NfL levels in subjects who carried one or two *APOE4* alleles. The reported effects are subtle and should be verified by independent cohorts.

This study emphasizes the importance of studying the effects of *APOE4* homozygosity and heterozygosity separately, by demonstrating significant differences in white matter integrity between these groups and their association to other biomarkers for AD.

## Authors’ contribution

CT, AS and EP analyzed data for this study. CT and AS drafted the manuscript. AS, LE SH and RP contributed to data collection. JR, AS and LE conceptualized the study. VS, HZ and KB contributed to data analysis and interpretation. AS, LE and JR supervised the study. All authors read and critically revised the manuscript for its content and approved the final version.

## Data Availability

Data will be made available upon reasonable request to the authors

## Acknowledgments

The authors would like to acknowledge the study participants for their altruist contribution and the personal at Turku PET Centre for collecting the data for this study.

