## Supplementary figure 1 for "White matter integrity and its association with amyloid-PET and serum NfL in healthy *APOE4* homozygotes, heterozygotes and non-carries"

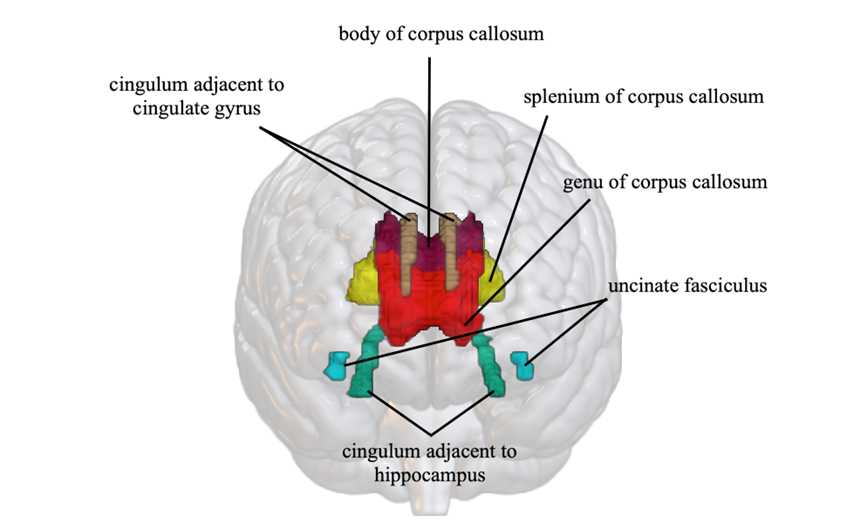

**Supplementary figure 1.** Regions of interest (ROIs): cingulum adjacent to cingulate gyrus (brown), body of corpus callosum (magenta), splenium of corpus callosum (yellow), genu of corpus callosum (red), uncinate fasciculus (blue), cingulum adjacent to hippocampus (turquoise)
