## Supplementary table 1 for "White matter integrity and its association with amyloid-PET and serum NfL in healthy *APOE4* homozygotes, heterozygotes and non-carries"

**Supplementary table 1.** Demographics including *APOE2/3*

|  | ***APOE4/4*** | ***APOE4/3*** | ***APOE3/3*** | ***APOE2/3*** | **p-value^a^** |
| --- | --- | --- | --- | --- | --- |
| n | 20 | 39 | 37 | 5 |  |
| Age (y), mean (SD) | 69.0 (4.72) | 71.0 (5.04) | 71.2 (5.62) | 73.4 (1.63) | 0.28 |
| Sex (M/F), n (%) | 7 / 13 (35% / 65%) | 14 / 25 (36% / 64%) | 15 / 22 (41% / 59%) | 4 / 1 (80% / 20%) | 0.28 |
| Education, n (%) |  |  |  |  | 0.70 |
| Primary school | 6 (30%) | 11 (28%) | 13 (35%) | 0 (0%) |  |
| Middle or comprehensive school | 6 (30%) | 10 (26%) | 9 (24%) | 2 (40%) |  |
| High school | 6 (30%) | 7 (18%) | 9 (24%) | 2 (40%) |  |
| College or university | 2 (10%) | 11 (28%) | 6 (16%) | 1 (20%) |  |
| BMI (kg/m^2^), mean (SD) | 26.3 (4.17) | 26.4 (3.63) | 28.1 (5) | 29.2 (6.84) | 0.21 |
| Scanner (scanner1 / scanner2), n (%) | 13 / 7 (65% / 35%) | 35 / 4 (90% / 10%) | 25 / 12 (68% / 32%) | 5 / 0 (100% / 0%) | **0.035** |
| Fazekas score, median (IQR) | 1.15 (0.18-1.71) | 0.92 (0.00-1.30) | 0.92 (0.00-1.46) | 0.69 (0.00-0.75) | 0.66 |
| Serum NfL pg/ml, median (IQR) | 22.0 (15.7-30.1) | 19.6 (13.6-23.4) | 16.6 (14.0-19.7) | 16.1 (12.2-20.8) | 0.32 |
| [^11^C]PiB SUVr, median (IQR) | 2.53 (1.75-2.86)** | 1.82 (1.52-2.41)* | 1.54 (1.43-1.77)* | 1.66 (1.62-1.71) | **0.0024** |

*Note: ^a^P-value refers to overall difference among the groups. Categorical variables were analyzed with χ^2^ test and numerical variables were analyzed with one-way ANOVA or Kruskal-Wallis test, pair-wise differences compared to APOE3/3 carriers are shown with star symbols: * p < 0.05, ** p < 0.01*

*M = male, F = female, SUVr = standardized uptake value ratio, BMI = body mass index, H = high, L = low, scanner1 = Philips Ingenuity 3.0T TF PET-MR, scanner2 = Philips Ingenia 3.0 T systems*
