## Supplementary figure 2 for "White matter integrity and its association with amyloid-PET and serum NfL in healthy *APOE4* homozygotes, heterozygotes and non-carries"

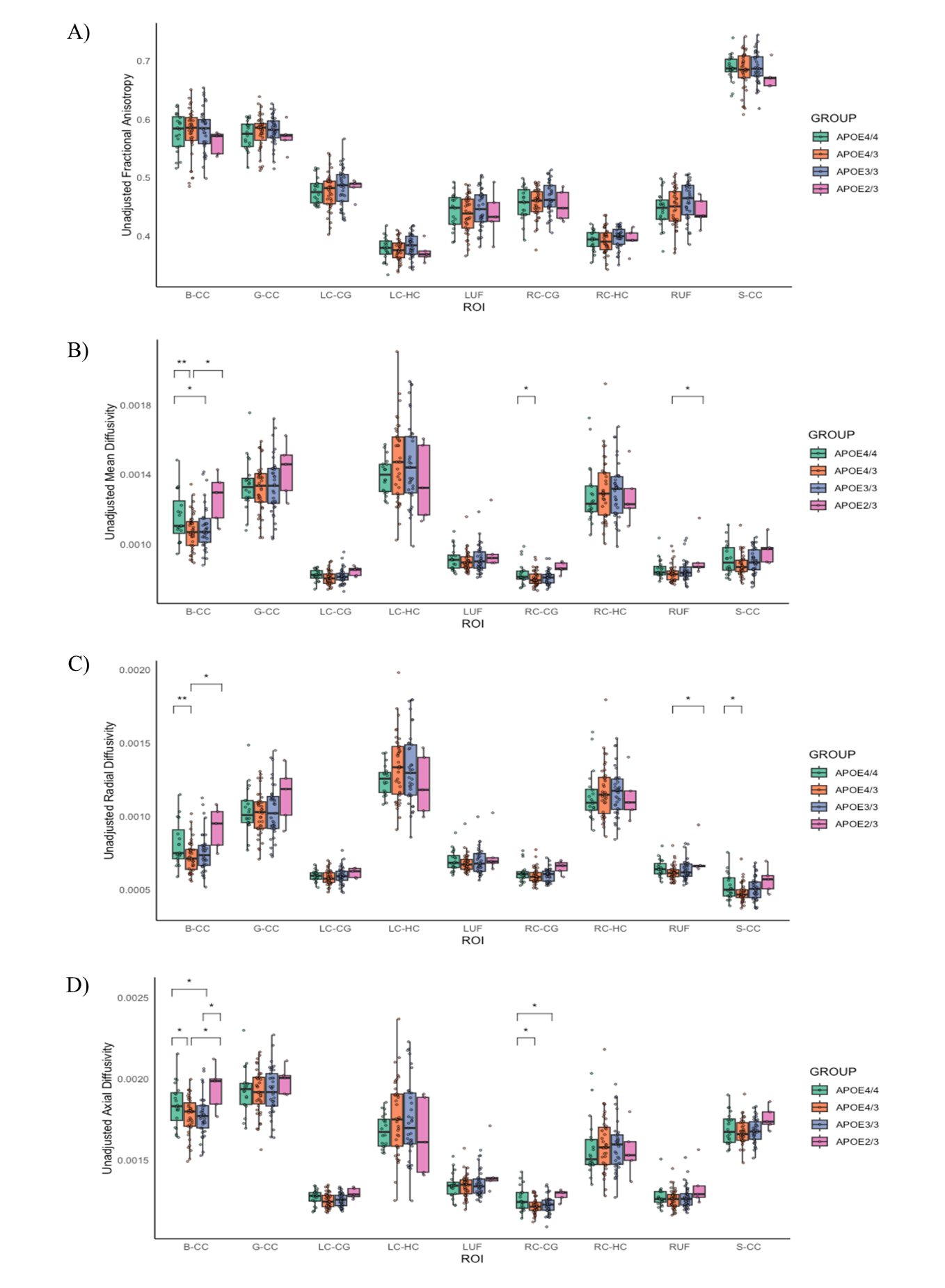
 **Supplementary figure 2.** Regional differences in fractional anisotropy (2A), mean diffusivity (2B), radial diffusivity (2C) and axial diffusivity (2D) between *APOE4/4*, *APOE4/3,* *APOE3/3* and *APOE2/3* estimated by ANCOVA, corrected for age and sex. Pair-wise differences in Tukey’s HSD tests are shown with star symbols: ** p < 0.05, ** p < 0.01*

B-CC = body of corpus callosum, G-CC = genu of corpus callosum, LC-CG = left cingulum adjacent to cingulate gyrus, LC-HC = left cingulum adjacent to hippocampus, LUF = left uncinate fasciculus, RC-CG = right cingulum adjacent to cingulate gyrus, RC-HC = right cingulum adjacent to hippocampus, RUF = right uncinate fasciculus, S-CC = splenium of corpus callosum
