## Supplementary figure 3 for "White matter integrity and its association with amyloid-PET and serum NfL in healthy *APOE4* homozygotes, heterozygotes and non-carries"

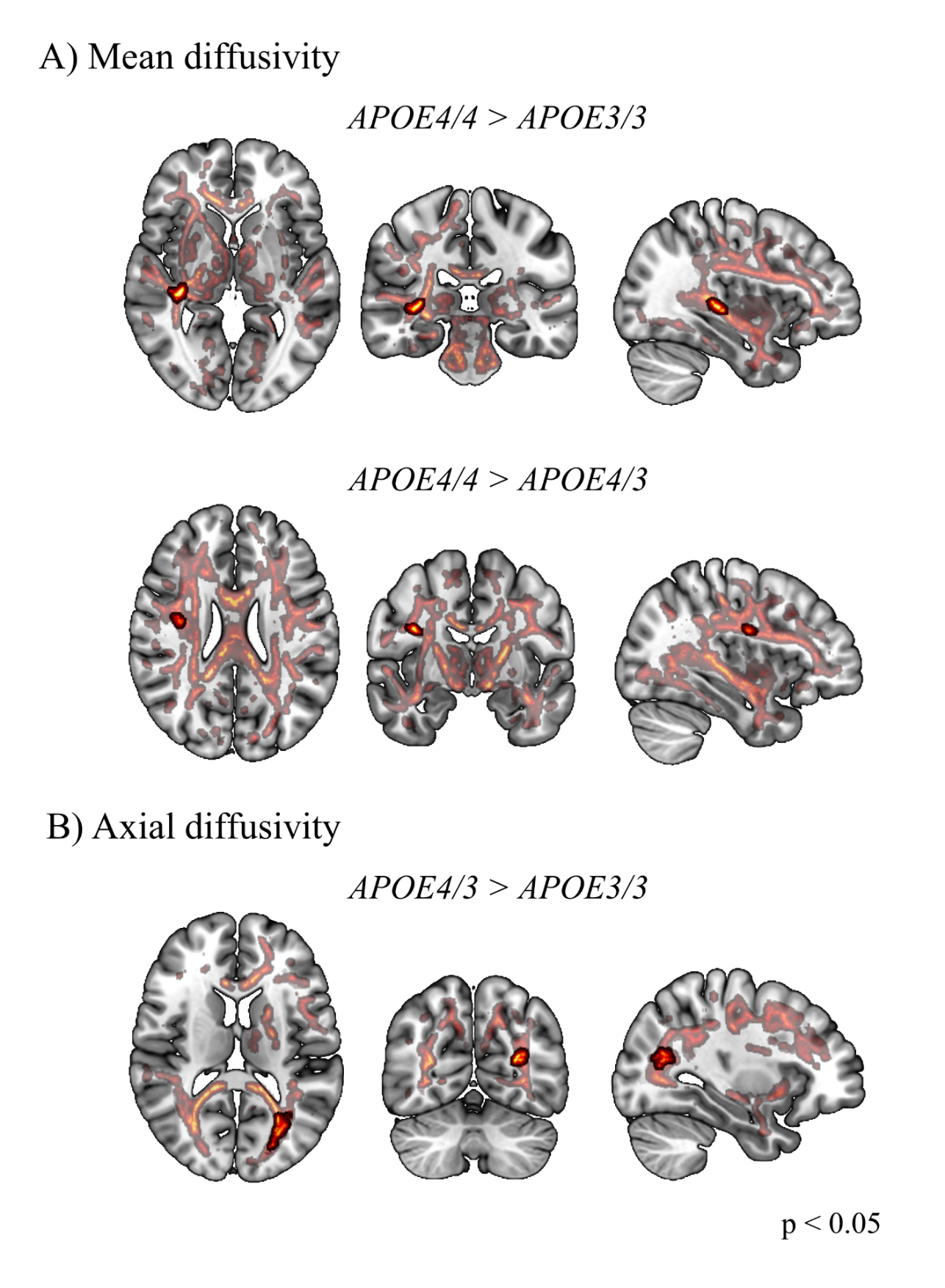

**Supplementary figure 3.** Significant clusters in TBSS analyses (shown in red) at p < 0.05, corrected for age and sex, uncorrected for multiple comparisons (semi-transparent) and controlling the family wise error rate with TFCE (opaque). *APOE4/4* show higher mean diffusivity than *APOE4/3* in the internal capsule and *APOE3/3* carriers in the right superior longitudinal fasciculus (3A); *APOE4/3* show greater axial diffusivity than *APOE3/3* in a cluster comprising part of the splenium of corpus callosum, left posterior thalamic radiation and left posterior corona radiata (3B).
